# Adding discharge characteristics to improve six-month post-discharge mortality prediction in under-five children with suspected sepsis in Ugandan hospitals

**DOI:** 10.64898/2026.03.27.26349094

**Authors:** Tanjila Akter, Nathan Kenya Mugisha, Vuong Nguyen, Abner Tagoola, Elias Kumbakumba, Hubert Wong, Jerome Kabakyenga, Niranjan Kissoon, Stephen Businge, J Mark Ansermino, Matthew O Wiens

**Author notes:** Corresponding author: Matthew O. Wiens. Institute for Global Health, BC Children’s Hospital, 305 - 4088 Cambie Street, Vancouver, BC, V5Z 2X8, Canada. Senior author (J Mark Ansermino, Matthew O Wiens).

## Abstract

**Background:** Many children under five die post hospital discharge in low-and middle-income countries (LMICs), particularly after treatment for severe infections. While some models exist, evidence on risk prediction for post-discharge mortality remains limited, with most relying solely on admission characteristics, overlooking in-hospital disease progression and discharge features.

**Methods:** We used secondary data from prospective cohort studies in six Ugandan hospitals (2012-2021) to update models at discharge. Of 8,810 children included, 3,665 were aged <6 months and 5,145 were aged 6-60 months. Models were developed utilizing an elastic net regression approach, with admission variables selected a priori and discharge variables selected based on variable importance ranking. Performance was evaluated by applying 10-fold cross-validation, area under the receiver operating characteristic curve (AUROC), Brier score, and Net Reclassification Index (NRI).

**Results:** Models augmented with discharge characteristics outperformed admission-only models. For children aged <6 months, the model AUROC improved by 5.1% (95% CI 3.0 – 7.3, *P<0.001*), achieving an AUROC of 0.81 and a Brier score of 0.06. In the 6–60m cohort, the model AUROC increased by 4.4% (95% CI 2.0 – 6.9, *P<0.001*), with an AUROC of 0.79 and a Brier score of 0.04. The NRI was 10.41% for children <6 months and 14.51% for those 6-60m and was achieved primarily through a reduction of false positive rates.

**Conclusion:** Adding only three discharge characteristics to the post-discharge mortality model based on admission characteristics enhanced prediction accuracy, including model calibration, discrimination and risk stratification compared to admission-only models.

**Key Messages:**

- Post-discharge mortality risk prediction models that incorporated discharge characteristics performed better than admission-only models for children under five in Uganda.
- The augmented model achieved stronger discrimination, improved calibration and substantial reclassification gains, primarily by reducing false positive rates.
- Most of the benefits of these improved models stem from accurately identifying low-risk survivors, which reduces unwarranted follow-up and enables the more effective utilization of the health system’s limited resources.

## Introduction

About five million under-5 deaths occur globally each year, most occurring in Sub-Saharan Africa and South Asia (1–3). Post-discharge mortality remains a significant yet underrecognized burden in LMICs, particularly in sepsis (4–7). Several recent studies have shown that as many children who are admitted to a health facility for acute illness or suspected sepsis die after discharge as during their initial hospitalization. Most of these deaths occur in the first few weeks following discharge, often at home or while in transit seeking care (8–10). Post-discharge mortality outcomes are also influenced by structural barriers to follow-up care, such as distance to facilities, transport costs, and household-level constraints. Therefore, innovative approaches to improving post-discharge outcomes are urgently required.

Early identification of children at increased risk of post-discharge mortality is a critical step towards building efficient and sustainable solutions to improve the transition of care from hospital to home(11,12). Clinical prediction models provide a data-driven method to identify children at high risk of post-discharge mortality, though these have not been integrated into routine care in most settings. The "Smart Discharge" program in Uganda has begun to tackle these challenges by integrating predictive algorithms, that utilize data collected during admission, to identify children at high risk of post-discharge mortality to target risk-specific interventions (12,13). These models are intended to support frontline clinical decision-making at discharge (e.g., nurses, clinical officers, and physicians) across regional referral and non-referral hospitals providing pediatric care.

Risk prediction at the time of admission allows healthcare providers to prioritize discharge and follow-up plans early during the hospitalization period. However, a child’s predicted risk on admission does not take into account the course of illness, response to therapy and any in-hospital complications and thus may not reflect the risk profile at discharge. Thus, re-evaluating risk at discharge to incorporate both admission and discharge characteristics is needed to identify high-risk children, to tailor follow-up plans, and enhance post-discharge care. Therefore, we aimed to develop models to update the admission predicted risk by integrating discharge-specific factors.

## Methods

### Study population and data source

This secondary analysis uses data from four previously published prospective cohorts that enrolled children with suspected sepsis from six hospitals in Uganda: Mbarara Regional Referral Hospital (Mbarara, southwest Uganda), Holy Innocents Children’s Hospital (Mbarara, southwest Uganda), Masaka Regional Referral Hospital (Masaka, central Uganda), Jinja Regional Referral Hospital (Jinja City, east Uganda), Villa Maria Hospital (central Uganda), and Uganda Martyrs Hospital, Ibanda (southwestern Uganda) between 2012 and 2021 (8,11,12). These hospitals serve urban and rural catchment populations across 30 districts and represent children under 5 years of age admitted with suspected sepsis in these settings. Children were enrolled at admission and followed for 6 months after discharge. Two cohorts included children aged 0–6 months, and two included children aged 6–60 months. Children living outside hospital catchment areas were excluded to reduce loss to follow-up for the post-discharge outcome. Additional details on eligibility criteria, follow-up procedures, sample size considerations, data collection, and approvals are provided in **Supplementary Text S1 (available as Supplementary data at *IJE* online).**

### Model development

#### Prior model overview (admission-only variable model)

The original models were developed to predict post-discharge mortality within six months of discharge using variables collected at the time of admission. These models were developed to be incorporated into a mobile application or an Electronic Health Record to identify high-risk patients at the time of hospital admission, thereby allowing discharge planning to occur throughout the hospitalization. Separate models were developed within two age groups (0-6 and 6-60 months) to ensure modelling would be possible under a variety of data availability scenarios. For the 0-6 months group, up to eight admission variables were incorporated, and for the 6-60 months group, nine variables were included, as the 9^th^ variable was reserved to reflect an interaction with another variable(12).

#### Updated model approach (updated discharge variable model)

To build upon prior admission-only variable models, we sought to generate a prediction update at the time of discharge through the incorporation of key discharge variables into the original model. Using this approach, the admission model remains in use to guide discharge planning; however, an updated risk score is generated at the time of discharge to inform further actions.

An intermediate model was first developed to explore variable importance and guide the selection of discharge predictors. Age interactions with all other predictors were included in this model to account for potential effect modification. The final model was then derived by reducing the intermediate model through variable importance and ranking.

For the under 6 months age group: **Model A** represents the admission-only variable model, **Model B** is the intermediate model with all admission and discharge variables, and **Model C** is the final reduced variable model. For the 6-60m age group: the corresponding models are **Model D** (admission-only)**, Model E** (intermediate) **and Model F** (final reduced variable model). Variables included in each model for both age cohorts can be found in the **Supplementary Table S1 (available as Supplementary data at *IJE* online).**

#### Outcome

The outcome of interest was mortality within 183 days of discharge.

#### Predictors

The predictors for the discharge variable models were of two categories. First, the admission predictors that were used in the original models were retained and used to ensure that the prior models would continue to provide clinical utility for discharge planning initiation. Second, 5 discharge features were prespecified a priori because they are routinely recorded at discharge across sites and plausibly reflect discharge readiness (physiologic stability, feeding function, and discharge disposition). These discharge variables included oxygen saturation, feeding status, discharge status (routine, referred or unplanned), respiratory rate and length of hospital stay (**Supplementary table S2, available as Supplementary data at *IJE* online).**

We also pre-specified that the final model would include a maximum of three discharge variables out of five initially considered. This decision was guided by prioritizing model parsimony and the feasibility of implementation in low-resource clinical settings.

#### Statistical analysis

We used the Elastic Net regression approach to develop and internally validate an improved set of risk prediction models. This robust Elastic Net technique combines the properties of the Least Absolute Shrinkage and Selection Operator (Lasso) and Ridge Regression, making it particularly advantageous for handling datasets with many correlated predictors(14). Firstly, we performed a univariate elastic net regression model to evaluate which predictors were associated with post-discharge mortality. Then, we developed full models using a total of 13 predictors, incorporating both admission and discharge characteristics for each age group. The tuning parameters- alpha and lambda were optimized using 10-fold cross-validation during model training, and optimal values were identified using grid search. All continuous predictors were mean-centred and scaled to unit variance, and categorical variables were transformed into dummy variables prior to the model development (**Supplementary Text S1)**.

We restricted our final model to have 11 predictors, eight previously identified admission predictors, regardless of their ranking in variable importance (VI) and the three top contributing discharge predictors. The final discharge predictors were selected by calculating and ranking VI in 10-fold cross-validation.

The model output was the predicted probability of six-month mortality. Performance was evaluated using the Brier score (overall accuracy), AUROC (with DeLong’s test comparing admission-only vs the final reduced variable models), and PR-AUC. Calibration was assessed using calibration plots. For model comparisons, we applied a prespecified probability threshold chosen to achieve 80% sensitivity in the admission-only model and generated ROC, PR, calibration, threshold, and lift plots. Internal validation used 10-fold cross-validation.

We compared models with vs without discharge predictors using the category-based net reclassification index (NRI), defining low vs high risk using the 80% sensitivity threshold (15). Missing data were handled using imputation; details are provided in **Supplementary Text S1 (available as Supplementary data at *IJE* online**). All analyses were conducted using R version 4.2.2(16).

## Result

### Study population and descriptive characteristics

A total of 8810 children were included:, 3665 in the under-6-month group and 5145 in the 6-60-month group. Six-month post-discharge mortality data were available for 3349 (97.8%) and 4830 (98.2%), respectively. Participant flow from enrolment to six-month outcome is shown in **Supplementary Figure S1**. Among children with available outcome data, 257 (7.7%) in the under-6-month cohort and 233 (4.8%) in the 6-60-month cohort died within six months after discharge. Descriptive statistics for the study population and candidate admission and discharge predictors are presented in **Table 1**. In the under-6-month cohort, 56.3% were male, the mean age was 2 months, and 59.3% had a duration of present illness of 48 hours to 7 days. In the 6–60-month cohort, 55.3% were male, the mean age was 22 months, and 6.6% had MUAC <115 mm. Median admission oxygen saturation was 96% in both cohorts. At discharge, 93.4% and 95.5% in the under-6-month and 6–60-month cohorts, respectively, were routinely discharged alive. Median length of stay was 5 (IQR 3–6) days and 4 (IQR 2–6) days, respectively. Univariate associations of admission and discharge predictors with six-month post-discharge mortality are shown in **Supplementary Table S3**. In both cohorts, adverse discharge conditions were strongly associated with post-discharge mortality.

**Table 1:** Characteristics of the study population and candidate admission and discharge predictors by age cohort.

| Characteristic | Under 6 months |  | 6–60 months |  |
| --- | --- | --- | --- | --- |
|  | Total N (%) / Mean (SD) / Median (Q1-Q3) | Missing, n (%) | Total N (%) / Mean (SD) / Median (Q1-Q3) | Missing, n (%) |
| Six-month outcome available, n (%) | 3349 (97.8%) |  | 4830 (98.2%) |  |
| Post-discharge mortality among those with outcome data, n (%) | 257 (7.7%) |  | 233 (4.8%) |  |
| Post-discharge survival among those with outcome data, n (%) | 3092 (92.3%) |  | 4597 (95.2%) |  |
| Age, months | 2 (1.9) | 0 (0%) | 22 (13.7) | 1 (0.02%) |
| Sex |  |  |  |  |
| Male | 1884 (56.3%) | 0 (0%) | 2670 (55.3%) | 0 (0%) |
| Female | 1465 (43.7%) |  | 2160 (44.7%) |  |
| MUAC, mm | 114 (18) | 3 (0.1%) | 139 (16) | 18 (0.4%) |
| <110 | 1304 (38.9%) |  | 321 (6.6%) |  |
| 110–120 | 942 (28.1%) |  | 514 (10.6%) |  |
| >120 | 1100 (32.8%) |  | 3977 (82.3%) |  |
| Weight-for-age z-score | -1.07 (1.97) | 2 (0.1%) | -1.29 (1.71) | 12 (0.2%) |
| < -3 | 463 (13.8%) |  | 668 (13.8%) |  |
| -3 to -2 | 356 (10.6%) |  | 723 (15.0%) |  |
| > -2 | 2528 (75.5%) |  | 3427 (71.0%) |  |
| SpO<sub>2</sub>, %<sup>+</sup> | 96 (92–99) | 9 (0.3%) | 96 (92–99) | 22 (0.5%) |
| <90% | 598 (17.9%) |  | 774 (16.0%) |  |
| 90% to 95% | 891 (26.6%) |  | 1236 (25.6%) |  |
| >95% | 1851 (55.3%) |  | 2798 (57.9%) |  |
|  | Total N (%) / Mean (SD) / Median (Q1-Q3) | Missing, n (%) | Total N (%) / Mean (SD) / Median (Q1-Q3) | Missing, n (%) |
| **HIV status** |  |  |  |  |
| HIV– | 3228 (96.4%) | 2 (0.1%) | 4664 (96.6%) | 22 (0.5%) |
| HIV+ | 119 (3.6%) |  | 144 (3.0%) |  |
| **How long since last admission** |  |  |  |  |
| Never | 2848 (85.0%) | 20 (0.6%) | 2647 (54.8%) | 20 (0.4%) |
| <7 days | 122 (3.6%) |  | 191 (4.0%) |  |
| 7 days to <1 month | 180 (5.4%) |  | 400 (8.3%) |  |
| 1 month to <1 year | 179 (5.3%) |  | 1175 (24.3%) |  |
| >1 year | 0 (0%) |  | 397 (8.2%) |  |
| **Abnormal BCS score** |  |  |  |  |
| No | 3064 (91.5%) | 0 (0%) | 4422 (91.6%) | 0 (0%) |
| Yes | 285 (8.5%) |  | 408 (8.4%) |  |
| **Respiratory rate, breaths/min** | 57.35 (17.0) | 5 (0.15%) | 48.1 (15.7) | 7 (0.1%) |
| **Temperature, °C** | 37.4 (0.92) | 1 (0%) | 37.7 (1.2) | 3 (0.1%) |
| <36.5 | 386 (11.5%) |  | 505 (10.5%) |  |
| 36.5 to 37.5 | 1699 (50.7%) |  | 1868 (38.7%) |  |
| 37.6 to 39.0 | 1072 (32.0%) |  | 1638 (33.9%) |  |
| >39.0 | 191 (5.7%) |  | 816 (16.9%) |  |
| **Duration of present illness (under 6 months only)** |  |  |  |  |
| <48 hours | 957 (28.6%) | 4 (0.1%) | — | — |
| 48 hours to 7 days | 1985 (59.3%) |  | — | — |
|  | Total N (%) / Mean (SD) / Median (Q1-Q3) | Missing, n (%) | Total N (%) / Mean (SD) / Median (Q1-Q3) | Missing, n (%) |
| 8 days to 1 month | 343 (10.2%) |  | — | — |
| >1 month | 60 (1.8%) |  | — | — |
| Neonatal jaundice (under 6 months only) |  |  |  |  |
| No | 3054 (91.2%) | 34 (1.0%) | — | — |
| Yes | 261 (7.8%) |  | — | — |
| Feeding well / sucking well (under 6 months only) |  |  |  |  |
| No | 1385 (41.4%) | 8 (0.2%) | — | — |
| Yes | 1956 (58.4%) |  | — | — |
| Abnormal fontanelle (under 6 months only) |  |  |  |  |
| No | 3211 (95.4%) | 6 (0.2%) | — | — |
| Yes | 132 (3.9%) |  | — | — |
| Discharge characteristics |  |  |  |  |
| Length of stay, *days*<sup>+</sup> | 5 (3–6) | 0 (0%) | 4 (2–6) | 0 (0%) |
| Discharge status |  |  |  |  |
| Referred to higher level of care | 164 (4.9%) | 2 (0.1%) | 101 (2.1%) | 0 (0%) |
| Routine discharge | 2810 (83.9%) |  | 4143 (85.8%) |  |
| Unplanned discharge | 373 (11.1%) |  | 586 (12.1%) |  |
| SpO<sub>2</sub> at discharge, %<sup>+</sup> | 97 (94–99) | 593 (17.7%) | 99 (96–99) | 813 (16.8%) |
| Feeding status at discharge |  |  |  |  |
| Feeding well | 3025 (90.3%) | 118 (3.5%) | 3673 (76.0%) | 836 (17.3%) |
| Feeding poorly | 171 (5.1%) |  | 295 (6.1%) |  |
|  | Total N (%) / Mean (SD) / Median (Q1-Q3) | Missing, n (%) | Total N (%) / Mean (SD) / Median (Q1-Q3) | Missing, n (%) |
| Not feeding at all | 35 (1.0%) |  | 27 (0.6%) |  |
| **Respiratory rate at discharge, breaths/min** | 48.99 (12.87) | 555 (16.6%) | 38.6 (9.5) | 1833 (37.9%) |

*Note: Variables marked with a “+” sign are non-normally distributed and are reported as Median (IQR) instead of Mean (SD)*

### 0-6-month discharge models

**Model B** had an AUROC of 0.82 (95% CI: 0.79–0.85), with a 10-fold cross-validation average of 0.81 (range: 0.74–0.86), PR-AUC was 0.37, and Brier score was 0.06. **Model B** identified weight-for-age z-score (VI: 99.85), referral to higher level of care (VI: 72.24), SpO2 at discharge (VI: 70.78), unplanned discharge (VI:52.58), and feeding poorly at discharge (VI: 52.15) had the highest variable importance scores **(Figure 1)**. Details on the model coefficients, variable importance rankings and the performance metrics in the cross-validation process are provided in **Supplementary tables S4-S6, available as Supplementary data at *IJE* online.**

**Figure 1:**
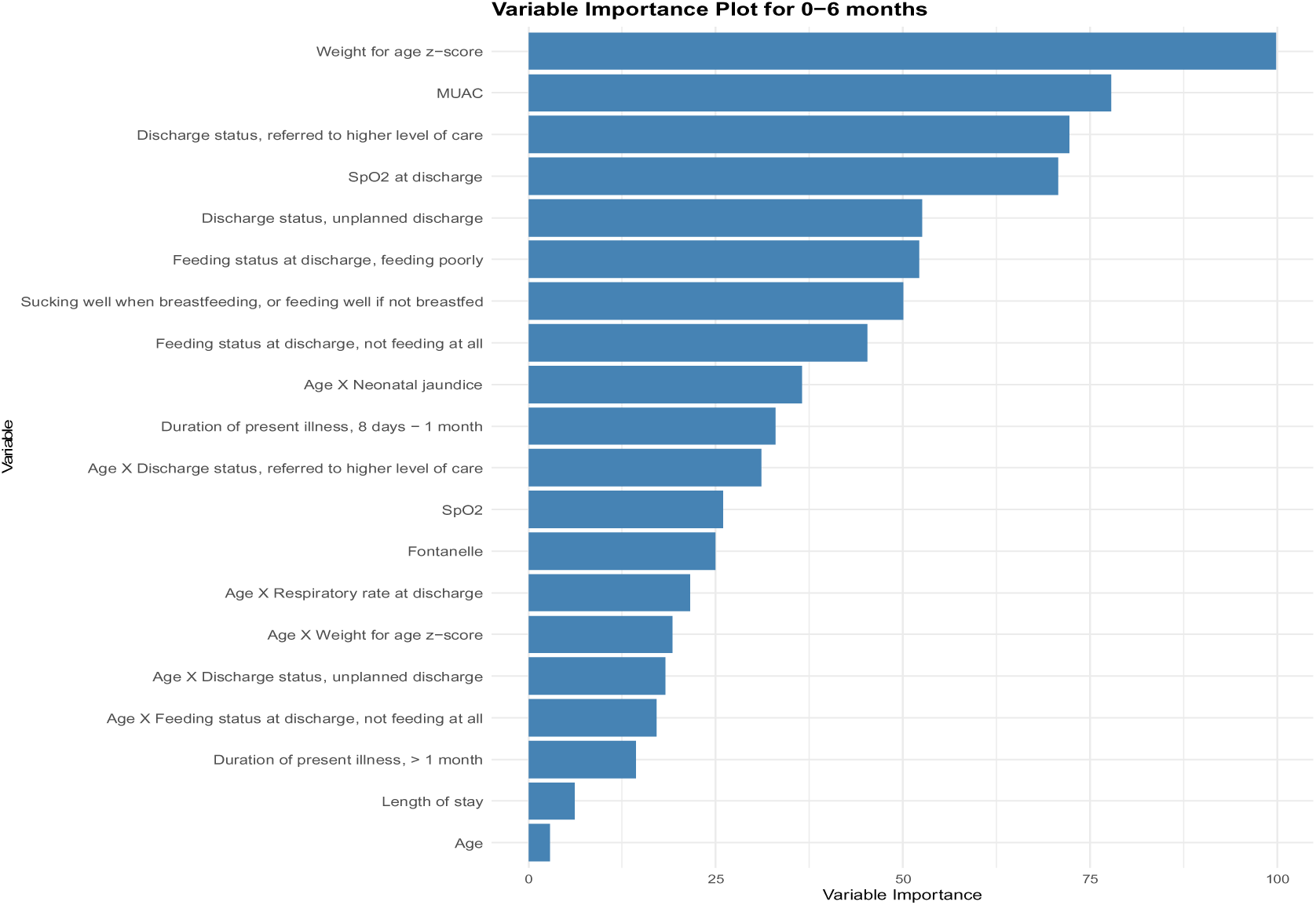
Variable importance plot for the intermediate model for the under-6 m cohort.

**Model C** was optimized with 11 key predictors, including three important discharge variables - SpO2 at discharge (mean rank[*r_m_*]= 4, selection frequency[*S_f_*]=10), Discharge status (*r_m_*=4, *S_f_* = 10), feeding status at discharge (*r_m_*=6, *S_f_* = 10)(**Supplementary table S11, available as Supplementary data at *IJE* online). Model C** maintained strong discriminatory power with an AUROC of 0.82 and the Brier score was 0.06 across multiple imputed datasets **(see Table 2)**. The model also performed well in 10-fold cross-validation, with a mean AUROC of 0.810 (range: 0.736–0.868), a mean PPV of 0.175 and a mean NPV of 0.974 with a mean PR-AUC of 0.353. Sensitivity was fixed at 80%, while specificity averaged 0.64 (**Supplementary table S12, available as Supplementary data at *IJE* online)**.

**Table 2:** Summary of performance for intermediate and final model using the probability threshold that gave a sensitivity of 0.8.

|  | *Under-6m cohort* |  | *6-60m cohort* |  |
| --- | --- | --- | --- | --- |
|  | **Model B** | **Model C** | **Model E** | **Model F** |
| **Full Dataset Performance** |  |  |  |  |
| Specificity | 0.66 | 0.68 | 0.69 | 0.67 |
| AUROC | 0.82 | 0.82 | 0.82 | 0.81 |
| PPV | 0.17 | 0.17 | 0.12 | 0.11 |
| NPV | 0.98 | 0.98 | 0.99 | 0.99 |
| PRAUC | 0.37 | 0.38 | 0.27 | 0.27 |
| Brier Score | 0.06 | 0.06 | 0.04 | 0.04 |
| **Average Cross-Validation Performance** |  |  |  |  |
| Specificity | 0.64 | 0.64 | 0.70 | 0.68 |
| AUROC | 0.81 | 0.81 | 0.80 | 0.79 |
| PPV | 0.17 | 0.18 | 0.14 | 0.14 |
| NPV | 0.97 | 0.97 | 0.98 | 0.98 |
| PRAUC | 0.35 | 0.35 | 0.25 | 0.26 |
| Brier Score | 0.06 | 0.06 | 0.04 | 0.04 |

**Note:** Model B =Intermediate model (0-6m), Model C =Final reduced model (0-6m), Model E=Intermediate model (6-60m), and Model F= Final reduced model (6-60m)

### 6-60 month discharge models

For 6-60m cohort, **Model E** showed a similar strong predictive performance, with an AUROC of 0.82 (95% CI: 0.79–0.85) and a 10-fold cross-validation average of 0.80 (range: 0.72–0.88). The PR-AUC was 0.27 (average: 0.25) and the Brier score was 0.04. Variable importance identified t SpO2 at discharge (VI: 76.40), unplanned discharge (VI: 73.74) and not feeding at all (VI: 40.97) as the top three discharge predictors (**Figure 2**). Details on model coefficients, variable importance rankings and the performance metrics in cross-validation process are provided in **Supplementary tables S7-S9, available as Supplementary data at *IJE* online.**

**Figure 2:**
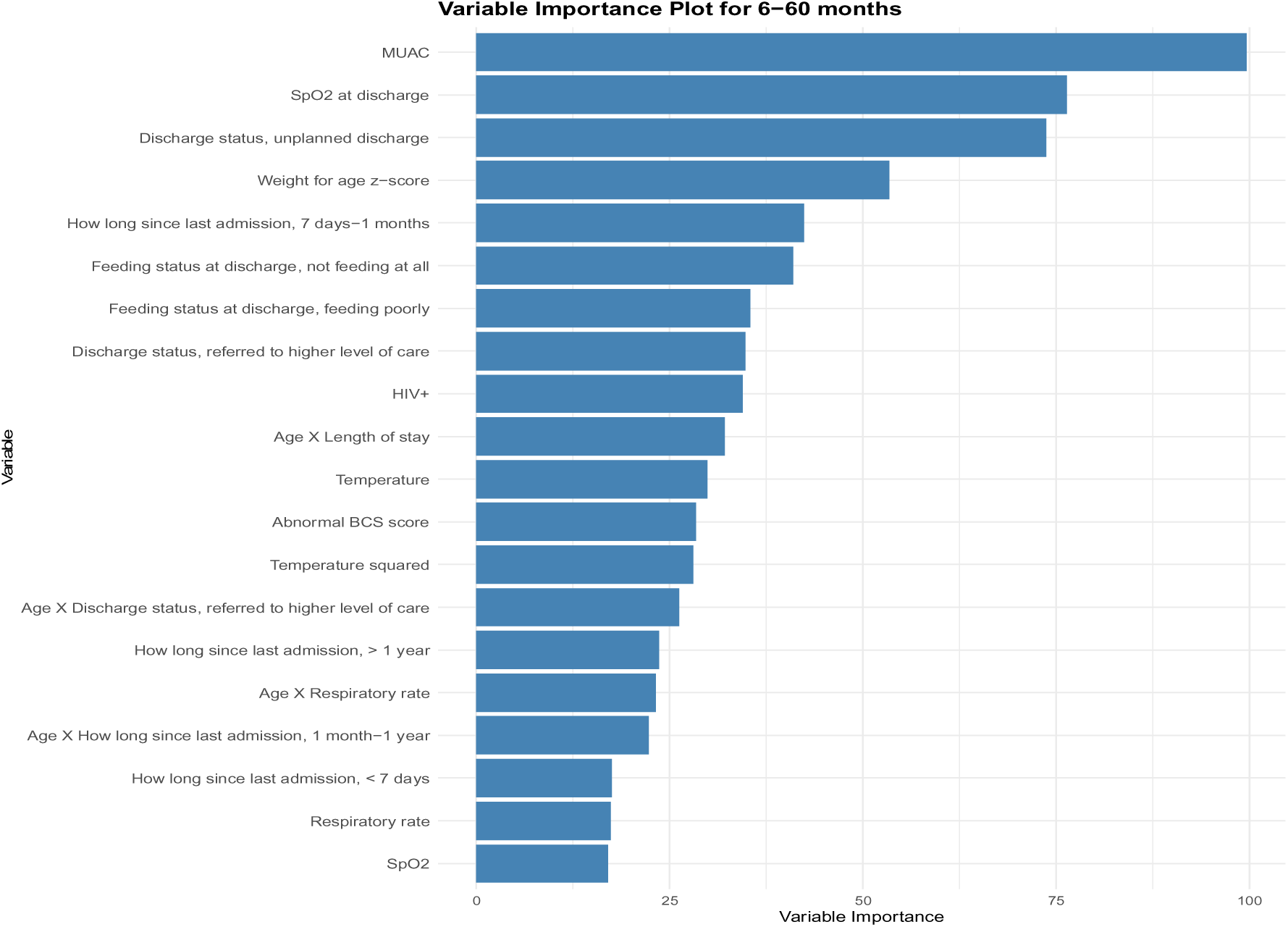
Variable importance plot for the intermediate model for the 6-60m cohort.

**Model F** was optimized using 12 key predictors, including three discharge variables - SpO2 at discharge (*r_m_*=2, *S_f_* = 10), discharge status (*r_m_*=3, *S_f_* = 10), and feeding status at discharge (*r_m_*=6, *S_f_* = 10) which were similar to those identified in **Model C** for the younger cohort (**Supplementary table S14, available as Supplementary data at *IJE* online). Model F** maintained high predictive accuracy (AUROC: 0.812, PR-AUC: 0.274 & Brier score: 0.04) across 17 imputed datasets (**see Table 2**). In 10-fold cross-validation, **Model F** achieved a mean AUROC of 0.791(range: 0.701 – 0.879), PPV of 0.136, and NPV of 0.983, with a PR-AUC of 0.268 and a Brier score of 0.041(**Supplementary table S15, available as Supplementary data at *IJE* online)**. The final model specifications, including intercepts and regression coefficients for individual risk calculation, are presented in **Supplementary tables S10 and S13, available as Supplementary data at *IJE* online.**

### Model comparison

#### Under-6-month cohort

**Model C** outperformed **Model A,** achieving an AUROC of 0.82 (95% CI: 0.79–0.85) compared to 0.77 (95% CI: 0.74–0.80); *P* <0.001 (DeLong test). The PR-AUC was 0.37 for **Model C** and 0.24 for **Model A**, and the PPV at 80% sensitivity was 0.17 and 0.13, respectively. Both models calibrated well at lower probabilities, however, **Model C** demonstrated superior calibration at higher risk levels (**Figure 3**).

**Figure 3:**
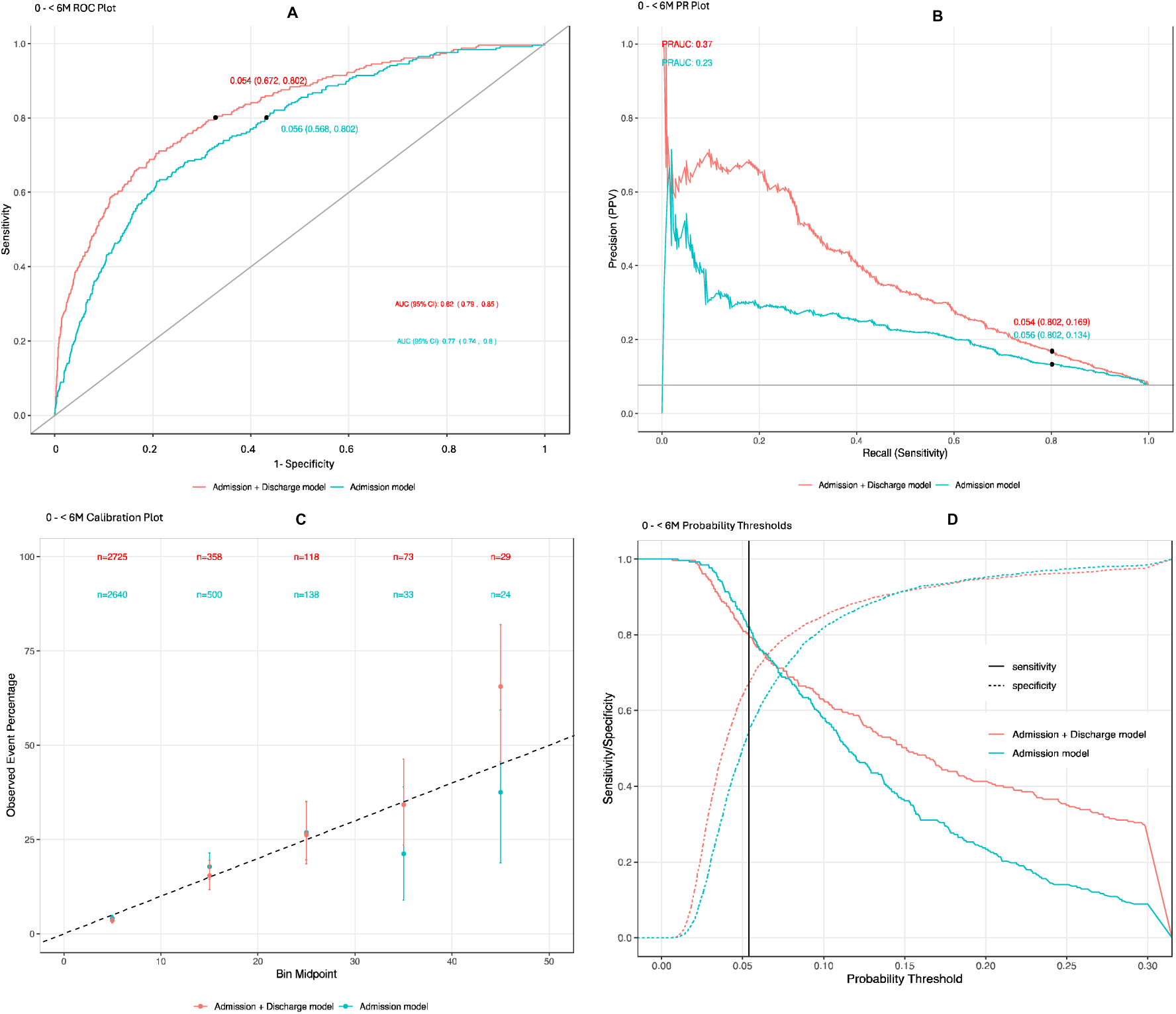
Model performance for predicting six-month post-discharge mortality in children aged under-6 m. (A) ROC curve (AUROC) comparing admission-only vs discharge-updated models. (B) Precision–recall curve (PR-AUC). (C) Binned calibration plot (observed vs predicted risk; dashed line indicates perfect calibration). (D) Sensitivity and specificity across probability thresholds; the vertical line indicates the prespecified threshold used for model comparison.

The lift chart highlighted greater efficiency in identifying mortality events. **Model C** identified 80% of events with 36% of the cohort sampled, while **Model A** required 46% to reach the same threshold (**See Supplementary Figure S1, available as Supplementary data at *IJE* online).**

#### 6–60-month cohort

Similarly, **Model F** showed superior performance (AUROC: 0.81, 95% CI: 0.78–0.84) than the **Model D (**AUROC: 0.75, 95% CI: 0.72–0.79); *P*=0.0004 (DeLong test). The PR-AUC was also higher (0.28 vs. 0.17), demonstrating better precision and recall. Calibration analysis confirmed **Model F** had a stronger alignment with observed outcomes across all risk levels (**Figure 4**).

**Figure 4:**
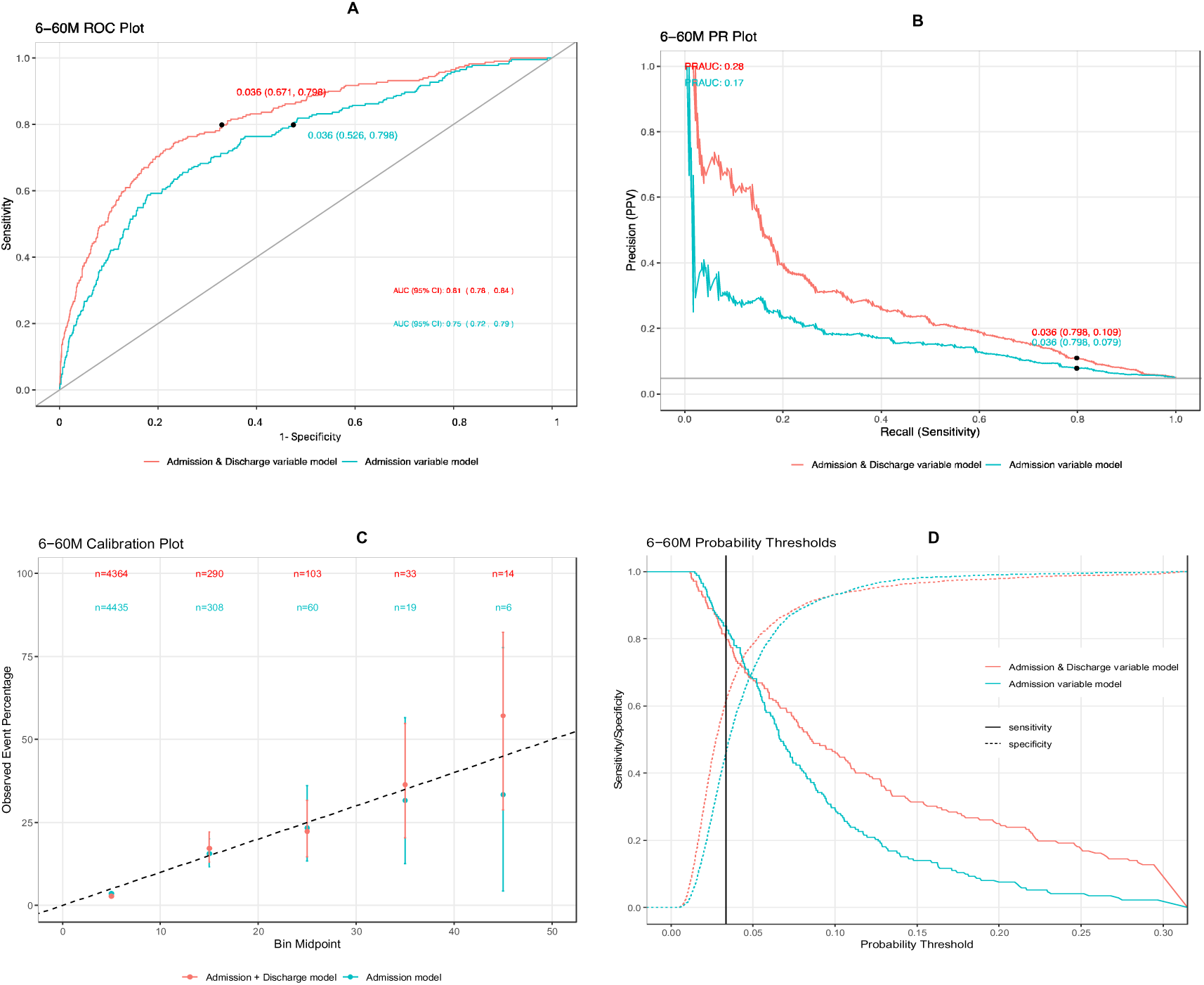
Model performance for predicting six-month post-discharge mortality in children aged 6-60 m. (A) ROC curve (AUROC) comparing admission-only vs discharge-updated models. (B) Precision–recall curve (PR-AUC). (C) Binned calibration plot (observed vs predicted risk; dashed line indicates perfect calibration). (D) Sensitivity and specificity across probability thresholds; the vertical line indicates the prespecified threshold used for model comparison.

The lift chart further supported this improvement, showing that at 35% of the cohort sampled, **Model F** identified 80% of events, compared to 72% by **Model D** (**See Supplementary Figure S1, available as Supplementary data at *IJE* online).**

### Net reclassification index (NRI)

At 80% sensitivity, the addition of discharge variables substantially improved risk classification particularly by reducing false positives. For the under-6-month cohort, the NRI was 10.41%, primarily driven by correctly reclassifying 14.97% of survivors from high to low risk (**See Table 3)** and for the 6–60-month group, 19.53% of survivors were correctly reclassified from high to low risk, yielding an overall NRI of 14.51% (**See Table 4**).

**Table 3:** Net reclassification index at the threshold where both models achieve 80% sensitivity for the under 6m cohort.

|  | Reclassified predicted risk (with discharge variables) |  | % (N) of subjects reclassified with |  |  |
| --- | --- | --- | --- | --- | --- |
| Predicted risk (without discharge variables) | Low | High | Increased risk | Decreased risk | Net correctly reclassified (%) |
| Post-discharge mortality=Yes, (N=257) |  |  |  |  |  |
| Low | 33 | 18 | 7.0% (18) | 7.0% (18) | 0 % |
| High | 18 | 188 |  |  |  |
| Post-discharge mortality=No, (N=3092) |  |  |  |  |  |
| Low | 1615 | 141 | 4.56% (141) | 14.97% (463) | 10.41 % |
| High | 463 | 873 |  |  |  |
| Net reclassification index | 10.41% |  |  |  |  |

**Footnote:** Low and high risk were defined using model-specific predicted-probability thresholds corresponding to approximately 80% sensitivity. For the under-6-month cohort, the threshold was 0.056 for the admission-only model and 0.054 for the admission + discharge model.

**Table 4:** Net reclassification index at the threshold where both models achieve 80% sensitivity for the 6-60 m cohort.

| Predicted risk (without discharge variables) | Reclassified predicted risk (with discharge variables) |  | % (N) of subjects reclassified with |  |  |
| --- | --- | --- | --- | --- | --- |
|  | Low | High | Increased risk | Decreased risk | Net correctly reclassified (%) |
| Post-discharge mortality=Yes, (N=233) |  |  |  |  |  |
| Low | 27 | 20 | 8.58% (20) | 8.58 % (20) | 0% |
| High | 20 | 166 |  |  |  |
| Post-discharge mortality=No, (N=4597) |  |  |  |  |  |
| Low | 2186 | 231 | 5.02 % (231) | 19.53% (898) | 14.51% |
| High | 898 | 1282 |  |  |  |
| Net reclassification index | 14.51% |  |  |  |  |

**Footnote:** Low and high risk were defined using model-specific predicted-probability thresholds corresponding to approximately 80% sensitivity. For the 6–60-month cohort, the threshold was 0.036 for both the admission-only model and the admission + discharge model.

## Discussion

We developed and internally validated a set of prediction models with improved discrimination and calibration for 6-month post-discharge mortality among Ugandan children under 5 years of age admitted with suspected sepsis, incorporating discharge variables, using four large prospective cohorts across six hospitals. This analysis highlights the importance of discharge characteristics in improving the risk stratification for post-discharge mortality. Compared with the original admission-only variable models, the updated prediction models at discharge consistently outperformed across both under-6-month and 6-60-month pediatric cohorts.

A strength of the updated model is its ability to provide a comprehensive risk assessment at discharge, addressing a key limitation of the previous model. The presence of adverse discharge features (such as feeding poorly or being hypoxic) among children who were being discharged is concerning and probably reflects a lack of discharge readiness. However, this discharge decision often reflects ward congestion, limited bed availability, and families’ pressing need to return home due to financial or caregiving responsibilities (17). Many discharges against medical advice (DAMA) cases arise from these factors, triggering a high-risk scenario requiring intentional follow-up.

Children needing specialized care are often referred to higher-level treatment, which follows a one-way trajectory from lower-level facilities to higher-level(18). Among children who were referred, our model flags those who have high post-discharge mortality risk scores at discharge, as they are more vulnerable and complex cases. These risk scores should accompany the child to the higher-level facility to ensure that the receiving facility’s medical staff is aware of the child’s risk profile. This enables a more data-driven approach to follow-up plans after the eventual discharge, since these high-risk cases may require more intensive follow-up after discharge than those referred to higher levels of care who do not meet the threshold for high risk. Similarly, our model precisely identifies children at high risk of post-discharge mortality among those who left against medical advice. These DAMA cases are challenging since they often result from socioeconomic limitations rather than clinical stability, which may lead to sub-optimal outcomes(19,20). By flagging those DAMA cases, our models enable the healthcare system to implement proactive follow-up strategies, including immediate phone call follow-ups and community health worker visits. These targeted interventions ensure continuity of care for DAMA cases, reducing the likelihood of negative consequences following early discharge.

As health systems begin to transition towards the use of risk algorithms to guide care, consideration must be given not only to model performance but also to practical considerations of model implementation such as the timing of implementation across the patient journey and actionable processes based on changes to predicted risk. In the context of high and persistent rates of pediatric post-discharge mortality, a concerted effort by health workers, policy makers and industry partners is necessary to develop strategies to identify and care for vulnerable children during the post-discharge period.

The Smart Discharges evidence-based risk prediction tool has shown the potential to save lives by reducing pediatric post-discharge mortality, evidenced by a recent trial in Uganda, where a risk-stratified discharge intervention reduced 6-month mortality from 6.0 % to 4.9 %(22). However, those discharge planning tools for risk-stratification are solely based on predictors collected during the admission period, which may not fully capture a child’s evolving risk profile during the hospitalization period. Incorporating discharge models to refine the risk score adds value, especially by reducing unnecessary follow-up or patient hospitalization costs(23). While implementing these models into routine care in resource-scarce environments poses practical logistical challenges (24), both the admission-only and expanded discharge models require similar effort to integrate into systems and, once introduced, will be well accepted by clinicians.

## Limitations

This study has several limitations. First, although our internal cross-validation results are promising, the model has not yet been externally validated in independent clinical settings which is essential to establishing the model’s generalizability and encouraging its adoption in various demographics and contexts of health systems. We have previously validated the admission-variables-only model in a Rwandan cohort(25) and plan to do the same for the new model. In addition, although the study included children from multiple hospitals serving both urban and rural catchment areas, the sample was restricted to Ugandan children admitted with suspected sepsis in participating hospital catchment areas, which may limit representativeness and generalizability to other settings. Second, another challenge remains the management of missing data, particularly in resource-limited settings where data collection is less standardized. While we used multiple imputation for model development which leverages the relationship between variables across the entire dataset, these techniques may not be feasible for point-of-care deployment due to their computational challenges. In practice, a lightweight imputation approach, such as mean/median imputation, may be more viable but needs to be validated (26). Finally, the updated model includes three discharge-specific features- discharge status, feeding status and Spo2-based pulse oximetry. While pulse oximetry may not be universally available in low-resource settings, expanding access and training for pulse oximetry use is now a priority in global health (27).

## Conclusion

This study presents an enhanced post-discharge mortality prediction model to identify children at risk of post-discharge death by integrating both the admission and discharge features in the model without substantially increasing the burden of data collection. In addition to improving prediction precision, the updated models provide a valuable tool for more effective resource allocation and care prioritization by reducing false positive rates. Future studies should focus on integrating this model digitally into the health system, streamlining workflows, and conducting external validation to ensure broader applicability.

## Ethics approval

This study involved a secondary analysis of a deidentified dataset. The parent study (Smart discharges to improve post-discharge health outcomes in children: A prospective before–after study with staggered implementation) was approved by the Mbarara University of Science and Technology (MUST) Research Ethics Committee (REC) (15/10-16, approved November 28, 2017), the Uganda National Council for Science and Technology (HS 2207, approved April 12, 2017), and the University of British Columbia/Children & Women’s Health Centre of British Columbia (UBC/C&W) Research Ethics Board (REB) (H16-02679, approved May 9, 2017). Written informed consent was obtained from parents or guardians of participating children in the parent study, and verbal assent was obtained from children where appropriate. As part of the consent process, participants were informed that their data would be deidentified and may be used for future research studies. The dataset used in this analysis was fully deidentified prior to secondary use. All procedures were conducted in accordance with the ethical standards of the MUST REC and UBC/C&W REB, and with the Helsinki Declaration of 1975.

## Author contributions

T.A. performed the study design, produced the analysis plan, conducted the statistical analysis, prepared the results, and wrote the first version of the manuscript. M.O.W. supervised the study, contributed to the study design, was involved in data preparation and data interpretation, and critically revised the manuscript. J.M.A. reviewed the analysis and provided substantive feedback on the manuscript. H.W. advised on the statistical methodology. V.N. and T.X. reviewed and provided feedback on the manuscript. N.K.M., A.T., E.K., J.K., N.K., and S.B. contributed to study oversight, data collection, and critical review of the final manuscript. All authors read and approved the final version of the manuscript. M.O.W. acts as the guarantor for the paper.

## Supplementary data

Supplementary data are available at IJE online.

## Data availability

The data underlying this article are available from the Smart Discharge Dataverse within the Borealis Dataverse repository. Access to the data can be granted upon request to the study investigators, in accordance with institutional data-sharing polices.

