## Supplemental materials for "Adding discharge characteristics to improve six-month post-discharge mortality prediction in under-five children with suspected sepsis in Ugandan hospitals"

Supplementary Information

Supplementary text S1

**Study population and data source**

This secondary analysis uses data from four previously published prospective cohorts that enrolled children with suspected sepsis from six hospitals in Uganda: Mbarara Regional Referral Hospital (Mbarara, southwest Uganda), Holy Innocents Children’s Hospital (Mbarara, southwest Uganda), Masaka Regional Referral Hospital (Masaka, central Uganda), Jinja Regional Referral Hospital (Jinja City, east Uganda), Villa Maria Hospital (central Uganda), and Uganda Martyrs Hospital, Ibanda (southwestern Uganda) between 2012 and 2021 (1–3). These facilities serve 30 districts (population~ 8.2 million; ~1.4 million under five), yielding a representative sample across urban and rural settings (ref). Briefly, children admitted with suspected sepsis were enrolled at admission and followed until six months post-hospital discharge. Of the four cohorts, two enrolled children from 0 to 6 months of age, while the other two enrolled children aged 6 to 60 months. All cohorts were enrolled by the same research staff, using identical inclusion criteria. Studies were approved by the institutional review boards of the Mbarara University of Science and Technology in Mbarara, Uganda (No. 15/10-16) and the University of British Columbia in Vancouver, Canada (H16-02679). Additionally, this study received approval from the Uganda National Council for Science and Technology (HS 2207).

Children were eligible if they were (1) admitted and (2) identified as having a proven or suspected infection by the treating medical team. Children were excluded if they lived outside the hospital's catchment area (to reduce loss to follow-up), were admitted for a brief observation period, or were admitted immediately after birth without first being discharged.

Study nurses collected clinical, socio-demographic, and baseline characteristics at admission and again recorded the discharge diagnosis, disposition, oxygen saturation, respiratory rate, feeding status, and length of hospital stay using encrypted study tablets at the point of discharge. A field officer ascertained six-month vital status using a standardized follow-up protocol: caregivers were contacted by telephone at 2 and 4 months after discharge and visited in person at 6 months to confirm survival status and, when applicable, record the date of death. Follow-up procedures were applied uniformly to all participants, irrespective of sociodemographic characteristics. Data were recorded on encrypted study tablets and uploaded to a REDCap database hosted at the BC Children’s Hospital Research Institute (Vancouver, Canada).

**Sample size**

The sample size for the primary study enrolment was determined to accomplish three primary aims. First, to explore the epidemiology of post-discharge mortality, which has been previously reported. Second, to develop prediction models. Third, to act as a control period for a later interventional phase.

For the present analysis, we determined the sample size required to develop a prediction model based on criteria proposed by Riley et al., 2020(4). For binary outcomes, three criteria are recommended based on: 1) reducing overfitting (caused by small sample sizes or too many candidate predictors relative to the sample size or number of events), defined by an expected shrinkage of predictor effects by ≤10%; 2) a small absolute difference of 0.05 in the apparent and adjusted Nagelkerke’s R2 value of the model, whereby the apparent R^2^ reflects the model performance in the same way that was used to develop the model and the adjusted R^2^ is an approximately unbiased estimate of the model fit;(5) and 3) estimating the outcome proportion to within ±5% precision. The estimated sample size required to satisfy the three criteria was 2,117 and 1,551 for the 0-6-month and 6-60-month cohorts, respectively.

We also created post hoc learning curves of the sample size used to develop the model versus the area under the receiver operating characteristic curve (AUROC) when tested against a 20% hold-out set using the full set of variable predictors (**Figure A i**n Supplementary S1 Text)(6). This involved building models using an increasing subset of the population (up to 80% of the total sample) and evaluating their performance against the hold-out set. For the 6-60-month learning curve, we started with the initial 1,242 children from 2012-2014, followed by the first 258 children recruited starting from July 2017 (n=1,500), and thereafter added groups of 500 consecutively recruited children until 80% (n=3,864) of the available population was reached. For the 0-6-month learning curve, we started with the first 500 children recruited starting from January 2018 and continued to add groups of 500 consecutively recruited children until 80% (n=2,679) of the available population was reached.

The performance of the 6-60-month derivation model increased with increasing sample size up to approximately 2,500 children after which the AUROC stabilized. The low performance of the model using only the initial 1,242 children may indicate model drift. The AUROC for the 0-6-month derivation model stabilized at approximately 1,000 children. This suggests that increasing our sample sizes beyond what we have currently collected would not result in further improvement of model performance.

**Learning Curve of Training Sample Size vs Performance**

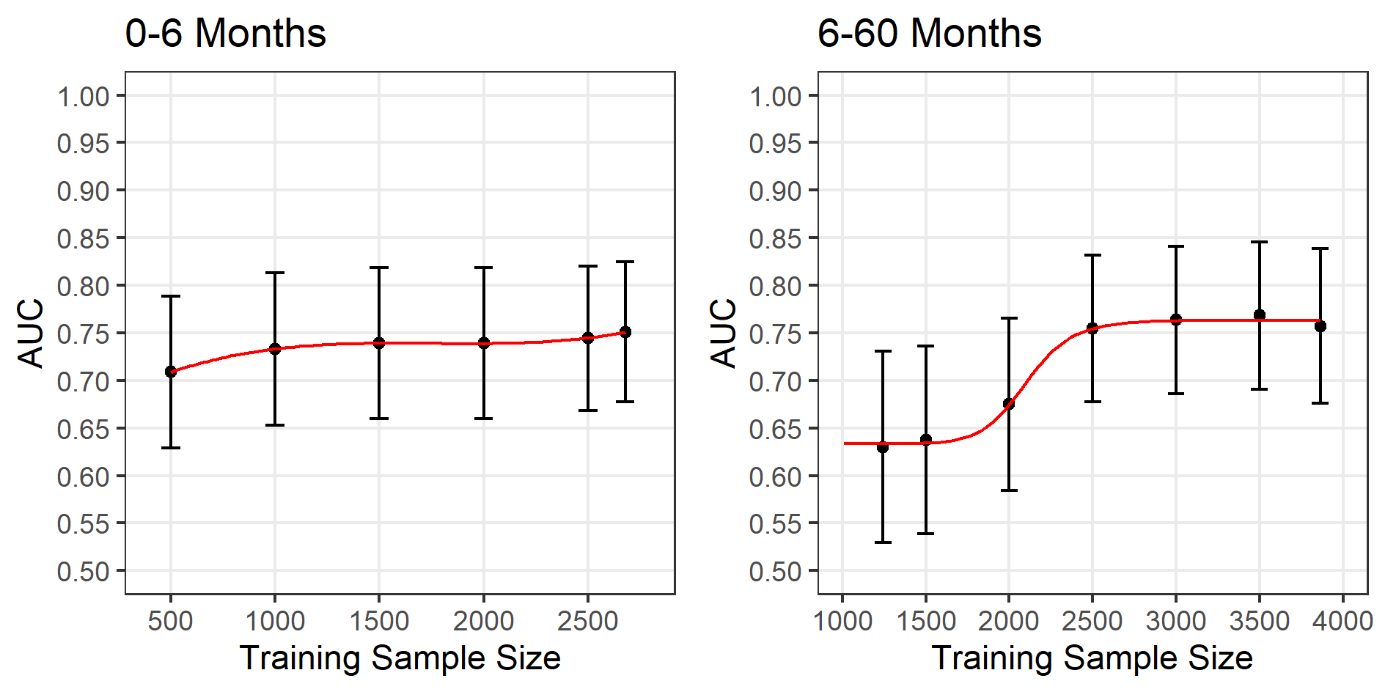

**Figure A.** Learning curve showing the performance in terms of area under the curve (AUC) and 95% confidence intervals of the elastic net derivation model against the training sample size.

**Adjustment and transformation of candidate predictors**

All continuous variables were mean-centred and scaled to unit variance (standardized to mean 0 and SD 1), and categorical variables converted to indicators (i.e. dummy variables). In addition to the raw oxygen saturation (SpO₂) measurement, we used the transformation for SpO₂ proposed by Zhou et al. to improve model prediction and calibration(7). Z-scored variable, weight-for-age z-scores and MUAC were calculated according to the World Health Organization (WHO) Child Growth Standards(8). We included a quadratic term for temperature since both high and low temperatures may increase risk. Other nonlinearities were not considered due to risk of overfitting with the limited number of available events per variable.

**Multiple imputation**

We imputed our admission dataset using K-nearest neighbour (KNN) to maintain consistency with our original admission-variables-only model (less than 5% of missingness). By contrast, discharge variables were more likely to be missing, with respiratory rate and oxygen saturation being absent in about 17% of cases. We used multiple imputation using chain equations (MICE) to address this(9,10), generating m=17 imputed datasets. We employed Predictive mean matching for numerical variables, Logistic regression imputation for binary variables, and Polytomous regression imputation for multicategory variables. Once 17 complete datasets were constructed using MICE, we fitted the elastic net regression model in each imputed dataset. Then, the estimated regression coefficients and their standard errors were extracted from the analysis conducted in each M-imputed dataset and combined using Rubin’s rules.

Supplementary table S1: List of models with predictors used for under 6 months and 6–60 months cohorts

| Model Name | Model Type | Variables |
| --- | --- | --- |
| Model A | ***Reference*** Model (Admission-variables only model) for under 6 months | Age, MUAC, SpO2; % at admission, Weight for age (z-score), Duration of present illness, Neonatal jaundice, Sucking well when breastfeeding/feeding, Fontanelle (total 8 predictors) |
| Model B | Full Unreduced ***Intermediate*** Model (Admission + discharge) for under 6 months | 8 Admission variables from $\boldsymbol{A}_{\boldsymbol{reference (0-<6)}}$ model & 5 additional discharge variables **(**SpO2, % at discharge, respiratory rate, bpm at discharge, feeding status at discharge, length of hospital stays, Discharge status of the children) |
| Model C | ***Final*** Model (Admission + discharge) for under 6 months | Reduced model with top 11 predictors selected based on variable ranking |
| Model D | ***Reference*** Model (Admission Variables) for 6-60 months | Age, MUAC, SpO2; % at admission, Weight for age (z-score), HIV status, how long since last admission, Abnormal BCS, Respiratory rate; bpm, Temperature; $℃$ (total 9 predictors) |
| Model E | Full Unreduced ***Intermediate*** Model (Admission + discharge) for 6-60 months | 9 Admission variables from $\boldsymbol{A}_{\boldsymbol{reference (6-60)}}$ model & 5 additional discharge variables **(**SpO2, % at discharge, respiratory rate, bpm at discharge, feeding status at discharge, length of hospital stays, Discharge status of the children) |
| Model F | ***Final*** Model (Admission + discharge) for 6-60 months | Reduced model with top 11 predictors selected based on variable ranking |

Supplementary table S2: Variables collected at admission and discharge point

| Variables | Applicable cohort | Definition/measurement | Categories |
| --- | --- | --- | --- |
| Age, months | Both | At admission, age from caregiver report | Continuous (month) |
| Duration of present illness | Under-6 only | **At admission**, time from symptom onset to presentation (caregiver report) | <48 hours, 48 hours to 7 days, 8 days to 1 month,  >1 months |
| MUAC (mm) | Both | **At admission**, MUAC measured using standard MUAC tape. | Continuous (month) |
| Neonatal jaundice | Under-6 only | **At admission**, jaundice present based on clinical assessment and/or caregiver report. | Binary: 1=present, 0=absent. |
| Sucking well when breastfeeding/feeding, binary | Under-6 only | **At admission**, feeding effectiveness assessed by observation and/or caregiver report. | Binary: 1=feeding well, 0=not well. |
| SpO2, % | Both | **At admission**, peripheral oxygen saturation measured by pulse oximeter. | Continuous (%). |
| Weight for age, z-score (WAZ) | Both | **At admission**, WAZ calculated from measured weight and age/sex using WHO standards. | Continuous (z-score). |
| Fontanelle, binary | Under-6 only | **At admission**, abnormal anterior fontanelle on examination. | Binary: 1=abnormal, 0=normal. (Define abnormal: bulging/sunken/etc.) |
| HIV, binary | 6-60 only | **During admission**, HIV status from documented result and/or admission testing per protocol. | Binary: 1=positive, 0=negative. |
| How long since last admission | 6-60 only | **At admission**, time since last hospital admission from caregiver report and/or record. | Never, <7 days, 7 days to <1 month, 1 month to <1 year, ≥1 year) |
| Abnormal BCS, binary | 6-60 only | **At admission**, Blantyre Coma Scale assessed clinically. | Binary: 1=abnormal, 0=normal. |
| Respiratory rate, bpm | 6-60 only | **At admission**, respiratory rate measured using RRate app by nurse. | Continuous (breaths/min). |
| Temperature, $\boldsymbol{℃}$ (<36.5, 36.5 to 37.5, 37.6 to 39, >39) | 6-60 only | **At admission**, temperature measured | <36.5; 36.5–37.5; 37.6–39.0; >39.0. |
| SpO2, % at discharge | Both | **At discharge**, pulse oximetry value closest to discharge (final assessment). | Continuous (%) |
| Respiratory Rate, bpm at discharge | Both | **At discharge**, respiratory rate closest to discharge (final assessment). | Continuous (breaths/min). |
| Feeding status, categorical at discharge  (feeding well,  feeding poorly, not feeding at all) | Both | **At discharge**, feeding status recorded by clinician/staff. | Feeding well; Feeding poorly; Not feeding at all. |
| Length of hospital stays, days | Both | From admission to discharge based on recorded dates/times. | Continuous (days). |
| Discharge status of the children, categorical (referred to higher level of care, routine discharge, unplanned discharge) | Both | **At discharge**, disposition recorded in the chart/study form. | Referred; Routine discharge; Unplanned discharge. |

Supplementary table S3: Univariate associations of admission and discharge predictors with post-discharge mortality.

*Children under 6 months and 6-60 months*

| **Variable** | **Under 6m OR (95% CI)** | **P-value** | **6-60m OR (95% CI)** | **P-value** |
| --- | --- | --- | --- | --- |
| **Admission predictors** | | | | |
| Age, months | 1.05 (0.98, 1.12) | 0.188 | 1.00 (0.99, 1.01) | 0.471 |
| **Sex** |  |  |  |  |
| Male | 1.18 (0.91, 1.53) | 0.218 | 0.90 (0.69, 1.17) | 0.433 |
| Female (ref.) | ref. |  | ref. |  |
| **MUAC, mm** | 0.96 (0.96, 0.97) | <0.001 | 0.96 (0.95, 0.97) | <0.001 |
| <110 | 3.81 (2.70, 5.51) | <0.001 | 6.66 (4.76, 9.25) | <0.001 |
| 110-120 | 1.58 (1.04, 2.42) | 0.033 | 2.77 (1.92, 3.92) | <0.001 |
| >120 (ref.) | ref. |  | ref. |  |
| **Weight-for-age z-score** | 0.71 (0.67, 0.75) | <0.001 | 0.71 (0.66, 0.76) | <0.001 |
| < -3 | 6.15 (4.58, 8.26) | <0.001 | 4.58 (3.40, 6.17) | <0.001 |
| -3 to -2 | 3.61 (2.51, 5.14) | <0.001 | 1.77 (1.20, 2.55) | 0.003 |
| > -2 (ref.) | ref. |  | ref. |  |
| **SpO2 at admission, %** | 0.96 (0.95, 0.98) | <0.001 | 0.95 (0.94, 0.97) | <0.001 |
| <90% | 1.78 (1.31, 2.41) | <0.001 | 2.07 (1.49, 2.84) | <0.001 |
| 90% to 95% | 0.87 (0.62, 1.20) | 0.406 | 1.15 (0.82, 1.58) | 0.404 |
| >95% (ref.) | ref. |  | ref. |  |
| **HIV status** |  |  |  |  |
| HIV- (ref.) | ref. |  | ref. |  |
| HIV+ | 1.37 (0.70, 2.42) | 0.317 | 3.81 (2.31, 6.00) | <0.001 |
| **How long since last admission** |  |  |  |  |
| Never (ref.) | ref. | <0.001 | ref. | <0.001 |
| <7 days | 2.04 (1.12, 3.47) | 0.013 | 2.37 (1.34, 3.94) | 0.002 |
| 7 days to <1 month | 2.68 (1.71, 4.06) | <0.001 | 2.39 (1.60, 3.52) | <0.001 |
| 1 month to <1 year | 2.47 (1.56, 3.79) | <0.001 | 1.42 (1.03, 1.94) | 0.031 |
| >1 year | — |  | 0.50 (0.22, 0.97) | 0.06 |
| **Abnormal BCS score** |  |  |  |  |
| No (ref.) | ref. |  | ref. |  |
| Yes | 2.37 (1.64, 3.34) | <0.001 | 1.93 (1.30, 2.78) | 0.001 |
| Respiratory rate at admission, breaths/min | 1.00 (0.99, 1.01) | 0.875 | 1.01 (1.00, 1.02) | 0.003 |
| **Temperature** | 0.90 (0.78, 1.04) | 0.167 | 0.81 (0.72, 0.91) | <0.001 |
| <36.5 | 0.96 (0.62, 1.43) | 0.835 | 1.28 (0.84, 1.89) | 0.234 |
| 36.5 to 37.5 (ref.) | ref. | 0.581 | ref. | 0.014 |
| 37.6 to 39.0 | 1.01 (0.76, 1.34) | 0.923 | 0.82 (0.60, 1.12) | 0.222 |
| >39.0 | 0.65 (0.32, 1.20) | 0.202 | 0.58 (0.37, 0.89) | 0.016 |
| **Duration of present illness (under 6 months only)** |  |  |  |  |
| <48 hours (ref.) | ref. | <0.001 | — | — |
| 48 hours to 7 days | 1.46 (1.05, 2.06) | 0.026 | — | — |
| 8 days to 1 month | 3.16 (2.09, 4.80) | <0.001 | — | — |
| >1 month | 5.13 (2.52, 9.87) | <0.001 | — | — |
| **Neonatal jaundice (under 6 months only)** |  |  |  |  |
| No (ref.) | ref. |  | — | — |
| Yes | 1.31 (0.83, 1.99) | 0.219 | — | — |
| **Sucking well when breastfeeding/feeding well if not breastfed (under 6 months only)** |  |  |  |  |
| No (ref.) | ref. |  | — | — |
| Yes | 0.47 (0.36, 0.61) | <0.001 | — | — |
| **Fontanelle (under 6 months only)** |  |  |  |  |
| No (ref.) | ref. |  | — | — |
| Yes | 2.40 (1.44, 3.81) | <0.001 | — | — |
| **Discharge predictors** | | | | |
| **Length of stay, days** | 1.06 (1.03, 1.08) | <0.001 | 1.01 (1.00, 1.02) | 0.003 |
| **Discharge status** |  |  |  |  |
| Referred to higher level of care | 10.12 (6.99, 14.54) | <0.001 | 11.48 (7.09, 18.15) | <0.001 |
| Routine discharge (ref.) | ref. | <0.001 | ref. | <0.001 |
| Unplanned discharge | 4.04 (2.92, 5.54) | <0.001 | 4.12 (3.04, 5.56) | <0.001 |
| **SpO2 at discharge, %** | 0.92 (0.90, 0.94) | <0.001 | 0.91 (0.89, 0.93) | <0.001 |
| **Feeding status at discharge** |  |  |  |  |
| Feeding well (ref.) | ref. | <0.001 | ref. | <0.001 |
| Feeding poorly | 5.30 (3.59, 7.70) | <0.001 | 4.37 (3.00, 6.25) | <0.001 |
| Not feeding at all | 19.30 (9.78, 38.72) | <0.001 | 15.07 (6.54, 32.90) | <0.001 |
| **Respiratory rate at discharge, breaths/min** | 1.02 (1.01, 1.03) | 0.001 | 1.02 (1.00, 1.04) | 0.017 |

***Note:*** ***Under-6-month-only predictors are shown as em dashes for the 6–60-month cohort. For non-binary categorical variables, the p-value for the reference group (labelled ref.) indicates the global p-value***

Supplementary table S4: Coefficient for full admission plus discharge variables model and variance for under-6 from different imputation datasets.

| Variable | Coefficient | SD |
| --- | --- | --- |
| (Intercept) | -2.857 | 0.067 |
| Age | 0.009 | 0.012 |
| Fontanelle | 0.073 | 0.017 |
| Neonatal jaundice | -0.011 | 0.018 |
| MUAC | -0.226 | 0.046 |
| Duration of present illness, 8 days – 1 month | 0.097 | 0.026 |
| Duration of present illness, > 1 month | 0.042 | 0.018 |
| SpO2 | -0.075 | 0.013 |
| Sucking well when breastfeeding, or feeding well if not breastfed | -0.146 | 0.034 |
| Weight for age z-score | -0.289 | 0.039 |
| Discharge status, referred to higher level of care | 0.209 | 0.025 |
| Discharge status, unplanned discharge | 0.153 | 0.026 |
| Feeding status at discharge, feeding poorly | 0.15 | 0.025 |
| Feeding status at discharge, not feeding at all | 0.129 | 0.01 |
| Length of stay | 0.017 | 0.009 |
| SpO2 at discharge | -0.204 | 0.04 |
| Respiratory rate at discharge | -0.011 | 0.021 |
| Age X Neonatal jaundice | 0.106 | 0.021 |
| Age X Duration of present illness, 48 hours-7 days | 0.031 | 0.029 |
| Age X Weight for age z-score | -0.05 | 0.033 |
| Age X Discharge status, referred to higher level of care | 0.088 | 0.012 |
| Age X Discharge status, unplanned discharge | 0.052 | 0.012 |
| Age X Feeding status at discharge, feeding poorly | -0.029 | 0.029 |
| Age X Feeding status at discharge, not feeding at all | 0.049 | 0.01 |
| Age X Respiratory rate at discharge | 0.066 | 0.041 |

Supplementary table S5: Average rank of variable importance and the number of times selected in the top 10 variables across 10 folds of cross-validation for full admissions plus discharge variables model for the under 6m.

| Variable | Rank | Selected |
| --- | --- | --- |
| Weight for age z-score | 1 | 10 |
| MUAC | 2 | 10 |
| Discharge status, referred to higher level of care | 3 | 10 |
| SpO2 at discharge | 4 | 10 |
| Sucking well when breastfeeding, or feeding well if not breastfed | 6 | 10 |
| Discharge status, unplanned discharge | 6 | 10 |
| Feeding status at discharge, feeding poorly | 6 | 10 |
| Feeding status at discharge, not feeding at all | 9 | 9 |
| Age X Neonatal jaundice | 9 | 8 |
| Duration of present illness, 8 days – 1 month | 9 | 8 |
| Age X Discharge status, referred to higher level of care | 12 | 3 |
| Fontanelle | 13 | 0 |
| Age X Respiratory rate at discharge | 13 | 0 |
| SpO2 | 14 | 2 |
| Age X Feeding status at discharge, feeding poorly | 17 | 0 |
| Age X Discharge status, unplanned discharge | 17 | 0 |
| Duration of present illness, > 1 month | 17 | 0 |
| Age X Feeding status at discharge, not feeding at all | 18 | 0 |
| Age X Duration of present illness, 48 hours-7 days | 18 | 0 |
| Neonatal jaundice | 21 | 0 |

Supplementary table S6: Performance metrics across 10 folds of cross-validation for full admissions plus discharge variables model for the under 6m.

| Fold | Threshold | Spec | Sens | AUC | PPV | NPV | PRAUC | Brier  Score |
| --- | --- | --- | --- | --- | --- | --- | --- | --- |
| 1 | 0.044 | 0.583 | 0.808 | 0.777 | 0.141 | 0.973 | 0.285 | 0.064 |
| 2 | 0.093 | 0.832 | 0.808 | 0.865 | 0.289 | 0.981 | 0.460 | 0.055 |
| 3 | 0.043 | 0.538 | 0.808 | 0.794 | 0.128 | 0.971 | 0.355 | 0.060 |
| 4 | 0.054 | 0.676 | 0.800 | 0.813 | 0.167 | 0.977 | 0.345 | 0.058 |
| 5 | 0.035 | 0.406 | 0.808 | 0.784 | 0.103 | 0.962 | 0.390 | 0.059 |
| 6 | 0.085 | 0.795 | 0.800 | 0.844 | 0.240 | 0.980 | 0.380 | 0.058 |
| 7 | 0.046 | 0.566 | 0.808 | 0.772 | 0.136 | 0.972 | 0.279 | 0.065 |
| 8 | 0.041 | 0.56 | 0.808 | 0.739 | 0.134 | 0.972 | 0.191 | 0.072 |
| 9 | 0.056 | 0.713 | 0.800 | 0.841 | 0.184 | 0.978 | 0.378 | 0.057 |
| 10 | 0.069 | 0.756 | 0.808 | 0.86 | 0.219 | 0.979 | 0.447 | 0.058 |
| Average | 0.057 | 0.642 | 0.806 | 0.809 | 0.174 | 0.974 | 0.351 | 0.061 |

Supplementary table S7: Coefficient for full admission plus discharge variables model and SD for 6-60 months from different imputation datasets.

| Variable | Coefficient | SD |
| --- | --- | --- |
| (Intercept) | -3.43 | 0.008 |
| Abnormal BCS score | 0.097 | 0.009 |
| HIV+ | 0.119 | 0.006 |
| MUAC | -0.343 | 0.008 |
| How long since last admission, < 7 days | 0.061 | 0.009 |
| How long since last admission, 7 days-1 months | 0.145 | 0.005 |
| How long since last admission, 1 month-1 year | 0.052 | 0.006 |
| How long since last admission, > 1 year | -0.081 | 0.005 |
| Respiratory rate | 0.059 | 0.009 |
| SpO2 | -0.06 | 0.02 |
| Temperature | -0.103 | 0.004 |
| Temperature squared | -0.097 | 0.004 |
| Weight for age z-score | -0.183 | 0.009 |
| Discharge status, referred to higher level of care | 0.119 | 0.008 |
| Discharge status, unplanned discharge | 0.253 | 0.012 |
| Feeding status at discharge, feeding poorly | 0.122 | 0.02 |
| Feeding status at discharge, not feeding at all | 0.143 | 0.023 |
| SpO2 at discharge | -0.259 | 0.037 |
| Respiratory rate at discharge | 0.016 | 0.03 |
| Abnormal BCS score X Age | -0.051 | 0.008 |
| Age X HIV+ | -0.009 | 0.008 |
| Age X How long since last admission, <7 days | 0.012 | 0.008 |
| Age X How long since last admission, 1 month-1 year | 0.077 | 0.006 |
| Age X Respiratory rate | 0.079 | 0.014 |
| Age X Discharge status, referred to higher level of care | 0.09 | 0.009 |
| Age X Feeding status at discharge, feeding poorly | 0.05 | 0.018 |
| Age X Feeding status at discharge, not feeding at all | 0.008 | 0.013 |
| Age X Length of stay | 0.111 | 0.003 |
| Age X Respiratory rate at discharge | 0.029 | 0.023 |

Supplementary table S8: Average rank of variable importance and the number of times selected in the top 10 variables across 10 folds of cross-validation for full admissions plus discharge variables model for the 6-60m.

| Variable | Rank | Selected |
| --- | --- | --- |
| MUAC | 1 | 10 |
| SpO2 at discharge | 2 | 10 |
| Discharge status, unplanned discharge | 3 | 10 |
| Weight for age z-score | 4 | 10 |
| Feeding status at discharge, not feeding at all | 6 | 10 |
| How long since last admission, 7 days-1 months | 7 | 10 |
| Feeding status at discharge, feeding poorly | 8 | 8 |
| HIV+ | 8 | 8 |
| Discharge status, referred to higher level of care | 9 | 6 |
| Age X Length of stay | 10 | 6 |
| Temperature | 12 | 3 |
| Age X Discharge status, referred to higher level of care | 13 | 2 |
| Temperature squared | 14 | 0 |
| Abnormal BCS score | 14 | 3 |
| How long since last admission, > 1 year | 16 | 1 |
| Age X How long since last admission, 1 month-1 year | 16 | 1 |
| Age X Respiratory rate | 18 | 1 |
| SpO2 | 18 | 1 |
| Respiratory rate | 19 | 0 |
| How long since last admission, < 7 days | 20 | 0 |

Supplementary table S9: Performance metrics across 10 folds of cross-validation for full admissions plus discharge variables model for the 6-60 m.

| Fold | Threshold | Spec | Sens | AUC | PPV | NPV | PRAUC | Brier  Score |
| --- | --- | --- | --- | --- | --- | --- | --- | --- |
| 1 | 0.025 | 0.474 | 0.783 | 0.736 | 0.069 | 0.978 | 0.254 | 0.041 |
| 2 | 0.045 | 0.741 | 0.792 | 0.788 | 0.142 | 0.985 | 0.231 | 0.043 |
| 3 | 0.058 | 0.831 | 0.783 | 0.837 | 0.190 | 0.987 | 0.277 | 0.040 |
| 4 | 0.065 | 0.878 | 0.783 | 0.881 | 0.246 | 0.988 | 0.365 | 0.038 |
| 5 | 0.059 | 0.848 | 0.783 | 0.871 | 0.207 | 0.987 | 0.300 | 0.039 |
| 6 | 0.033 | 0.663 | 0.783 | 0.722 | 0.104 | 0.984 | 0.195 | 0.043 |
| 7 | 0.030 | 0.508 | 0.783 | 0.735 | 0.075 | 0.979 | 0.152 | 0.044 |
| 8 | 0.033 | 0.612 | 0.783 | 0.774 | 0.094 | 0.982 | 0.302 | 0.04 |
| 9 | 0.053 | 0.805 | 0.792 | 0.849 | 0.177 | 0.987 | 0.287 | 0.041 |
| 10 | 0.035 | 0.636 | 0.792 | 0.785 | 0.103 | 0.983 | 0.165 | 0.046 |
| Average | 0.044 | 0.700 | 0.785 | 0.798 | 0.141 | 0.984 | 0.253 | 0.042 |

Supplementary table S10: Coefficient for final admission plus discharge variables model and SD for under-6 from different imputation datasets.

| Variable | Coefficient | SD |
| --- | --- | --- |
| (Intercept) | -2.936 | 0.027 |
| Age | 0.068 | 0.013 |
| Fontanelle | 0.097 | 0.008 |
| Neonatal jaundice | -0.046 | 0.019 |
| MUAC | -0.288 | 0.025 |
| Duration of present illness, 48 hours-7 days | 0.01 | 0.007 |
| Duration of present illness, 8 days – 1 month | 0.141 | 0.015 |
| Duration of present illness, > 1 month | 0.068 | 0.009 |
| SpO2 | -0.091 | 0.008 |
| Sucking well when breastfeeding, or feeding well if not breastfed | -0.194 | 0.018 |
| Weight for age z-score | -0.323 | 0.015 |
| Discharge status, referred to higher level of care | 0.223 | 0.009 |
| Discharge status, unplanned discharge | 0.172 | 0.015 |
| Feeding status at discharge, feeding poorly | 0.186 | 0.025 |
| Feeding status at discharge, not feeding at all | 0.137 | 0.014 |
| SpO2 at discharge | -0.226 | 0.034 |
| Age X Neonatal jaundice | 0.141 | 0.014 |
| Age X Duration of present illness, 48 hours-7 days | 0.088 | 0.017 |
| Age X Sucking well when breastfeeding, or feeding well if not breastfed | 0.021 | 0.011 |
| Age X Weight for age z-score | -0.021 | 0.021 |
| Age X Discharge status, referred to higher level of care | 0.1 | 0.01 |
| Age X Discharge status, unplanned discharge | 0.068 | 0.01 |
| Age X Feeding status at discharge, feeding poorly | -0.08 | 0.029 |
| Age X Feeding status at discharge, not feeding at all | 0.05 | 0.014 |
| Age X SpO2 at discharge | 0.02 | 0.013 |

Supplementary table S11: Average rank of variable importance and the number of times selected in the top 10 variables across 10 folds of cross-validation from the under for 6 m final admissions plus discharge variable models.

| Variable | Rank | Selected |
| --- | --- | --- |
| Weight for age z-score | 1 | 10 |
| MUAC | 2 | 10 |
| SpO2 at discharge | 4 | 10 |
| Discharge status, referred to higher level of care | 4 | 10 |
| Sucking well when breastfeeding, or feeding well if not breastfed | 5 | 10 |
| Feeding status at discharge, feeding poorly | 6 | 10 |
| Discharge status, unplanned discharge | 7 | 10 |
| Duration of present illness, 8 days – 1 month | 9 | 10 |
| Age X Neonatal jaundice | 9 | 8 |
| Feeding status at discharge, not feeding at all | 10 | 9 |
| Age X Discharge status, referred to higher level of care | 12 | 1 |
| Fontanelle | 13 | 0 |
| Age X Duration of present illness, 48 hours-7 days | 14 | 1 |
| SpO2 | 14 | 1 |

Supplementary table S12: Summary of performance across 10-fold cross-validation for the final admission plus discharge variables model for under 6 months using the probability threshold that gave a sensitivity of 0.80.

| Fold | Threshold | Spec | Sens | AUC | PPV | NPV | PRAUC | Brier  Score |
| --- | --- | --- | --- | --- | --- | --- | --- | --- |
| 1 | 0.045 | 0.599 | 0.808 | 0.779 | 0.146 | 0.974 | 0.288 | 0.064 |
| 2 | 0.095 | 0.833 | 0.808 | 0.868 | 0.29 | 0.981 | 0.455 | 0.055 |
| 3 | 0.041 | 0.536 | 0.808 | 0.795 | 0.128 | 0.971 | 0.365 | 0.06 |
| 4 | 0.053 | 0.688 | 0.8 | 0.812 | 0.172 | 0.977 | 0.346 | 0.057 |
| 5 | 0.032 | 0.382 | 0.808 | 0.783 | 0.099 | 0.959 | 0.391 | 0.059 |
| 6 | 0.084 | 0.794 | 0.8 | 0.85 | 0.239 | 0.98 | 0.382 | 0.058 |
| 7 | 0.044 | 0.548 | 0.808 | 0.766 | 0.131 | 0.971 | 0.283 | 0.065 |
| 8 | 0.04 | 0.547 | 0.808 | 0.736 | 0.13 | 0.971 | 0.189 | 0.072 |
| 9 | 0.057 | 0.717 | 0.8 | 0.842 | 0.186 | 0.978 | 0.381 | 0.057 |
| 10 | 0.067 | 0.769 | 0.808 | 0.865 | 0.228 | 0.979 | 0.452 | 0.056 |
| Average | 0.056 | 0.641 | 0.805 | 0.81 | 0.175 | 0.974 | 0.353 | 0.06 |

Supplementary table S13: Coefficient for final admission plus discharge variables model and variance for 6-60m from different imputation datasets.

| Variable | Coefficient | SD |
| --- | --- | --- |
| (Intercept) | -3.419 | 0.008 |
| Abnormal BCS score | 0.095 | 0.009 |
| HIV+ | 0.122 | 0.006 |
| MUAC | -0.349 | 0.008 |
| How long since last admission, < 7 days | 0.063 | 0.009 |
| How long since last admission, 7 days-1 months | 0.144 | 0.005 |
| How long since last admission, 1 month-1 year | 0.05 | 0.005 |
| How long since last admission, > 1 year | -0.079 | 0.007 |
| Respiratory rate | 0.049 | 0.007 |
| SpO2 | -0.064 | 0.02 |
| Temperature | -0.105 | 0.004 |
| Temperature squared | -0.099 | 0.004 |
| Weight for age z-score | -0.198 | 0.008 |
| Discharge status, referred to higher level of care | 0.118 | 0.008 |
| Discharge status, unplanned discharge | 0.253 | 0.012 |
| Feeding status at discharge, feeding poorly | 0.117 | 0.02 |
| Feeding status at discharge, not feeding at all | 0.14 | 0.023 |
| SpO2 at discharge | -0.26 | 0.034 |
| Abnormal BCS score X Age | -0.041 | 0.008 |
| Age X How long since last admission, <7 days | 0.013 | 0.008 |
| Age X How long since last admission, 1 month-1 year | 0.079 | 0.005 |
| Age X Respiratory rate | 0.119 | 0.007 |
| Age X Discharge status, referred to higher level of care | 0.089 | 0.009 |
| Age X Feeding status at discharge, feeding poorly | 0.052 | 0.018 |

Supplementary table S14: Average rank of variable importance and the number of times selected in the top 10 variables across 10 folds of cross-validation from the 6-60 m final admissions plus discharge variable models.

| Variable | Rank | Selected |
| --- | --- | --- |
| MUAC | 1 | 10 |
| SpO2 at discharge | 2 | 10 |
| Discharge status, unplanned discharge | 3 | 10 |
| Weight for age z-score | 4 | 10 |
| How long since last admission, 7 days-1 months | 6 | 10 |
| Feeding status at discharge, not feeding at all | 6 | 10 |
| HIV+ | 8 | 8 |
| Discharge status, referred to higher level of care | 9 | 7 |
| Feeding status at discharge, feeding poorly | 9 | 7 |
| Age X Respiratory rate | 11 | 5 |
| Temperature | 11 | 4 |
| Temperature squared | 13 | 2 |
| Abnormal BCS score | 13 | 3 |
| Age X Discharge status, referred to higher level of care | 13 | 2 |
| Age X How long since last admission, 1 month-1 year | 15 | 1 |
| How long since last admission, > 1 year | 16 | 0 |
| SpO2 | 17 | 1 |

Supplementary table S15: Summary of performance across 10-fold cross-validation for the final admission plus discharge variables model for 6-60 months using the probability threshold that gave a sensitivity of 0.80.

| Fold | Threshold | Spec | Sens | AUC | PPV | NPV | PRAUC | Brier  Score |
| --- | --- | --- | --- | --- | --- | --- | --- | --- |
| 1 | 0.024 | 0.443 | 0.783 | 0.721 | 0.066 | 0.976 | 0.262 | 0.04 |
| 2 | 0.042 | 0.714 | 0.792 | 0.789 | 0.131 | 0.985 | 0.236 | 0.043 |
| 3 | 0.052 | 0.813 | 0.783 | 0.835 | 0.174 | 0.987 | 0.285 | 0.04 |
| 4 | 0.066 | 0.884 | 0.783 | 0.879 | 0.257 | 0.988 | 0.378 | 0.038 |
| 5 | 0.06 | 0.849 | 0.783 | 0.87 | 0.208 | 0.987 | 0.304 | 0.039 |
| 6 | 0.029 | 0.579 | 0.783 | 0.701 | 0.085 | 0.982 | 0.193 | 0.043 |
| 7 | 0.03 | 0.451 | 0.783 | 0.712 | 0.068 | 0.976 | 0.15 | 0.044 |
| 8 | 0.035 | 0.643 | 0.783 | 0.778 | 0.099 | 0.983 | 0.322 | 0.039 |
| 9 | 0.051 | 0.794 | 0.792 | 0.842 | 0.169 | 0.986 | 0.285 | 0.041 |
| 10 | 0.034 | 0.62 | 0.792 | 0.779 | 0.1 | 0.983 | 0.167 | 0.046 |
| Average | 0.042 | 0.679 | 0.785 | 0.791 | 0.136 | 0.983 | 0.258 | 0.041 |

Supplementary Figure S1: Participant flow from enrolment to six-month outcome.

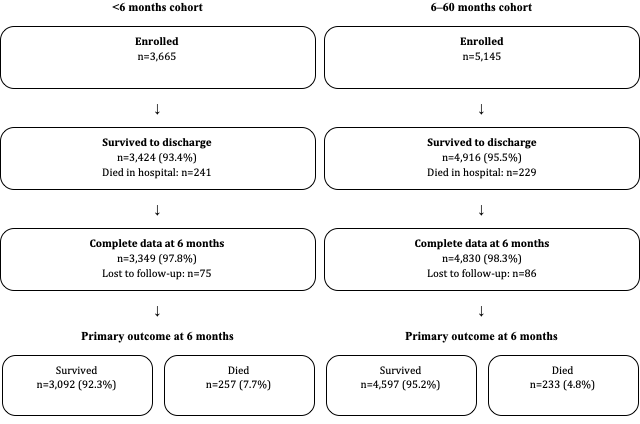

Supplementary figure S2: Lift chart comparison for under-6- and 6–60-month cohort.

B

A

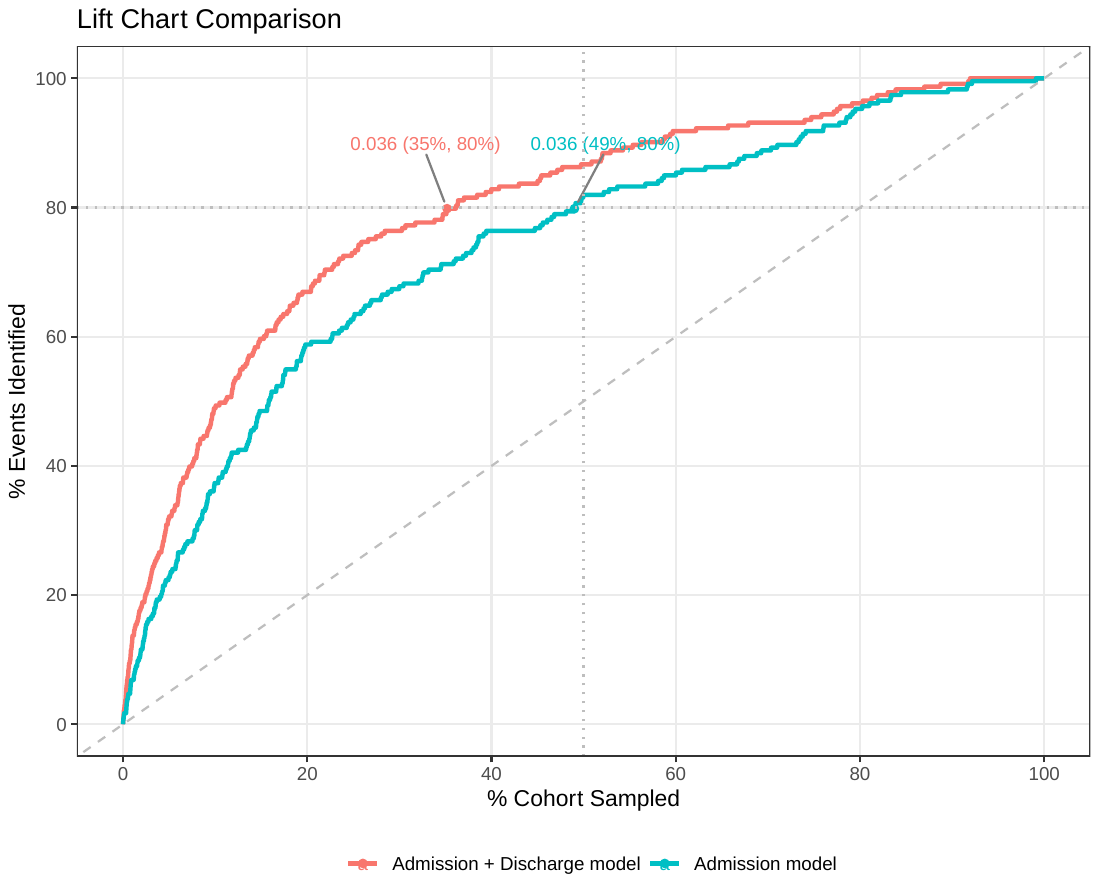

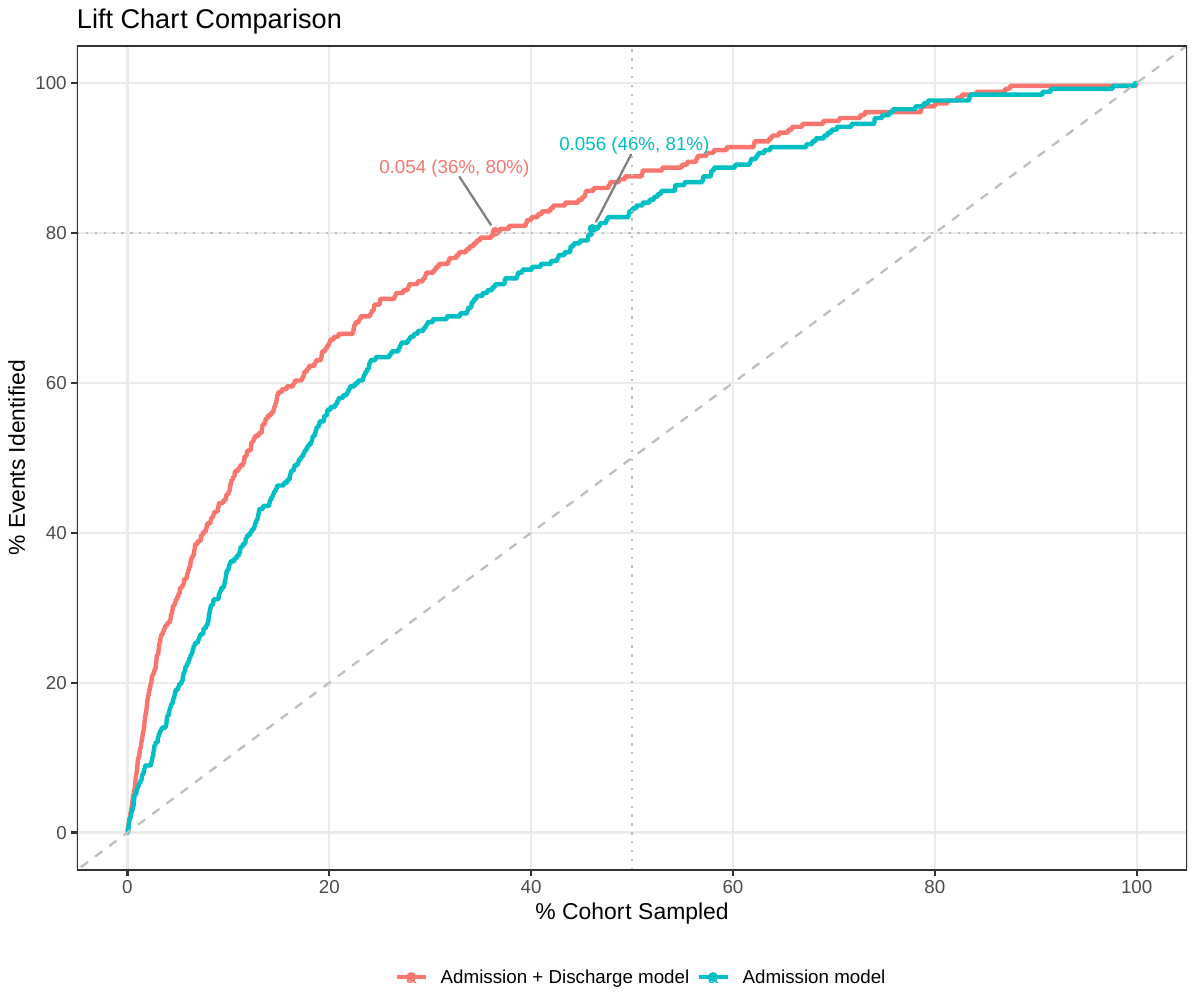
